# Expression quantitative trait methylation analysis elucidates gene regulatory effects of DNA methylation: The Framingham Heart Study

**DOI:** 10.1101/2022.04.13.22273839

**Authors:** Amena Keshawarz, Helena Bui, Roby Joehanes, Jiantao Ma, Chunyu Liu, Tianxiao Huan, Shih-Jen Hwang, Brandon Tejada, Meera Sooda, Paul Courchesne, Peter J. Munson, Cumhur Y. Demirkale, Chen Yao, Nancy L. Heard-Costa, Achilleas N. Pitsillides, Honghuan Lin, Ching-Ti Liu, Yuxuan Wang, Gina M. Peloso, Jessica Lundin, Jeffrey Haessler, Zhaohui Du, Michael Cho, Craig P. Hersh, Peter Castaldi, Laura M. Raffield, Jia Wen, Yun Li, Alexander P. Reiner, Mike Feolo, Nataliya Sharopova, Ramachandran S. Vasan, Edwin K. Silverman, Dawn L. DeMeo, April P. Carson, Charles Kooperberg, Daniel Levy

## Abstract

**Background:** Expression quantitative trait methylation (eQTM) analysis identifies DNA CpG sites at which methylation is associated with gene expression and may reveal molecular mechanisms of disease. The present study describes an eQTM resource of CpG-transcript pairs.

**Methods:** DNA methylation was measured in blood samples from 1,045 Framingham Heart Study (FHS) participants using the Illumina 450K BeadChip and in 1,070 FHS participants using the Illumina EPIC array. Blood gene expression data were collected from all 2,115 participants using RNA sequencing (RNA-seq). The association between DNA methylation and gene expression was quantified for all *cis* (i.e., within 1Mb) and *trans* (>1Mb) CpG-transcript pairs. Significant results (p<1E-7 for *cis* and <1E-14 for *trans*) were subsequently tested for enrichment of biological pathways and of clinical traits.

**Results:** We identified 70,047 significant *cis* CpG-transcript pairs where the top most significant eGenes (i.e., gene transcripts associated with a CpG) were enriched in biological pathways related to cell signaling, and for 1,208 clinical traits (enrichment false discovery rate [FDR] ≤ 0.05). We also identified 246,667 significant *trans* CpG-transcript pairs where the top most significant eGenes were enriched in biological pathways related to activation of the immune response, and for 1,191 clinical traits (enrichment FDR ≤ 0.05). Using significant *cis* CpG-transcript pairs, we identified significant mediation of the association between CpG sites and cardiometabolic traits through gene expression and identified shared genetic regulation between CpGs and transcripts associated with these cardiometabolic traits.

**Conclusions:** We developed a robust and powerful resource of eQTM CpG-transcript pairs that can help inform future functional studies that seek to understand the molecular basis of disease.

## Introduction

DNA methylation is an epigenetic modification characterized by the transfer of a methyl group onto DNA cytosine-phosphate-guanine (CpG) sites that can regulate gene expression. The extent of DNA methylation at specific CpG sites is associated with phenotypic variation in numerous traits including cardiovascular disease-related traits^1^ such as body mass index (BMI),^2^ blood lipids,^3^ glycemic traits,^4^ blood pressure,^5^ and inflammatory biomarkers.^6^ Expression quantitative trait methylation (eQTM) analysis identifies CpG sites that display methylation-related associations with expression of nearby (*cis*) or remote (*trans*) genes. While prior eQTM studies using commercial arrays for gene expression profiling have revealed associations between DNA methylation and gene expression that are linked to clinical disease, many of these studies are limited by small sample size and focus on specific disease phenotypes.^1,7–9^

To generate a resource of eQTM CpG-transcript pairs, we use array-based DNA methylation data and RNA sequencing (RNA-seq) gene expression data in over 2,000 Framingham Heart Study (FHS) participants to precisely examine the association between DNA methylation and gene expression. We posit that leveraging RNA-seq will increase the power, precision, and relevance of our eQTM analyses beyond what was possible with previous array-based gene expression studies.^7,10^ Use of RNA-seq gene expression data also provides an opportunity to interrogate DNA methylation in relation to long non-coding RNAs (lncRNAs) and explore their possible contributions to clinical disease.^11,12^

The primary aims of this study were to test the associations of DNA methylation with genome-wide gene expression and to create a resource of eQTM CpG-transcript pairs to facilitate new insights into disease pathways and processes. As proof of principle, we evaluated the associations of eQTM CpG-transcript pairs with cardiovascular traits and provide examples of how these eQTM resources can be used in future research.

## Methods

### Cohort description

An overview of the study design is depicted in **Figure 1**. DNA methylation and RNA-seq data were collected from 2,115 participants in the FHS Offspring (n=686) and Third Generation (n=1,429) cohorts. Peripheral whole blood samples were collected in the fasting state at the ninth examination cycle from FHS Offspring participants (2011-2014)^13^ and at the second examination cycle (2006-2009)^14^ from FHS Third Generation participants. All study protocols were approved by the Boston Medical Center Institutional Review Board, and all study participants provided their written informed consent.

**Figure 1.**
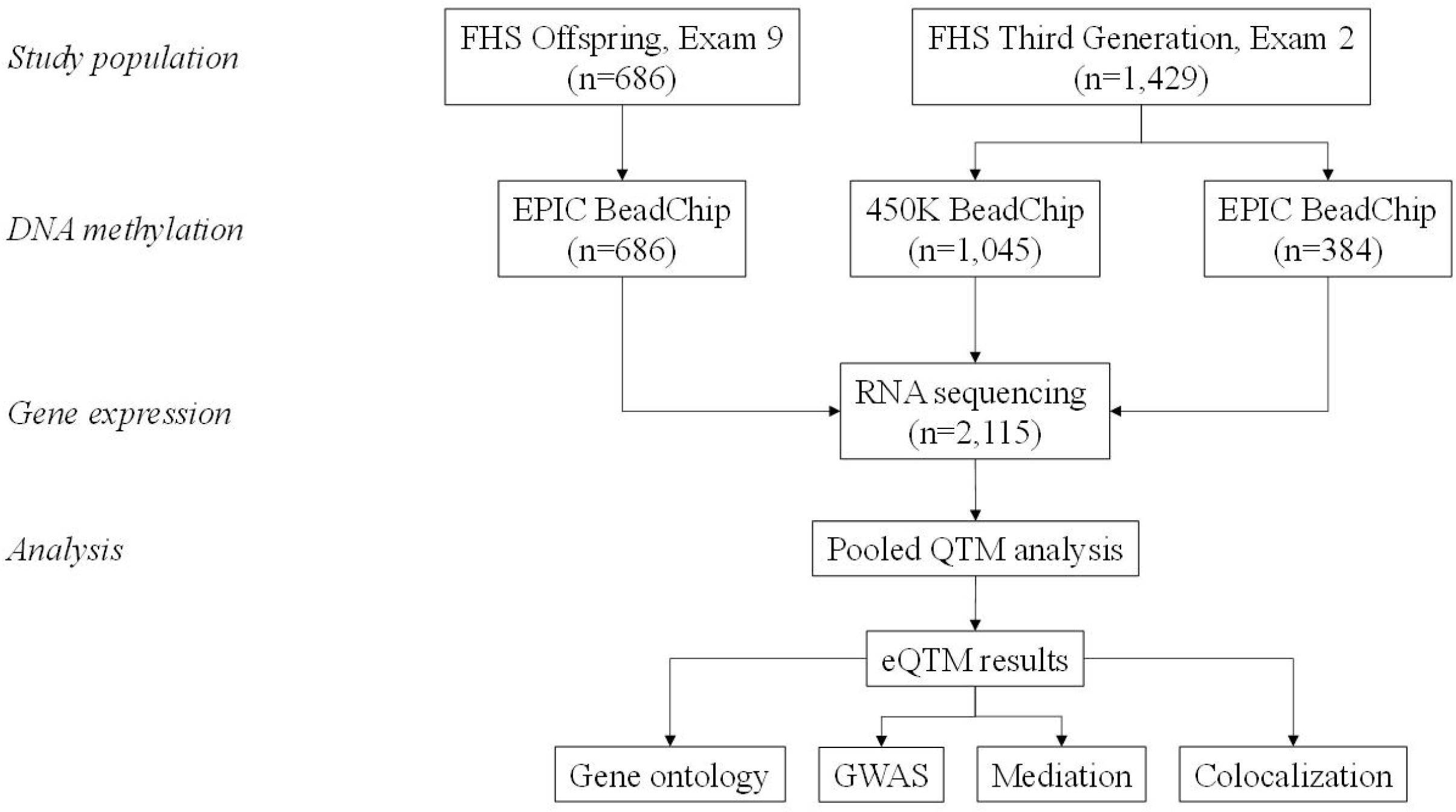
Overview of study design.

### DNA methylation data collection

Buffy coats were isolated from whole blood samples and prepared with bisulfite conversion for the DNA methylation assays. Samples from 1,045 FHS Third Generation participants were assayed for DNA methylation using the Infinium HumanMethylation450 BeadChip (Illumina Inc., San Diego, CA). Samples from the remaining 384 FHS Third Generation participants and all FHS Offspring participants were assayed for DNA methylation using the Infinium MethylationEPIC BeadChip (Illumina Inc., San Diego, CA). Methylation probes on autosomal chromosomes were analyzed while probes containing polymorphic SNPs were excluded.

### DNA methylation data pre-processing

DNA methylation data were pooled across both the HumanMethylation450 and the MethylationEPIC platforms, which shared 452,568 CpG loci. In addition to the data captured at these loci, samples assayed by the HumanMethylation450 platform had methylation data for 32,945 additional CpG sites, while those assayed by the MethylationEPIC platform had methylation data for 413,524 additional CpG sites.

Quality control (QC) and normalization were performed on the DNA methylation β values using the “dasen” function in the R *wateRmelon* package (version 1.16.0).^15^ Normalized β values were then residualized after accounting for batch effects, row effects, column effects, and four PCs constructed from the normalized β values. These residualized β values were subsequently winsorized at the mean ± 3 * standard deviation (SD) of the distribution for each probe.

Ten principal components (PCs) representing technical confounders were identified from the winsorized residuals. These ten PCs and the winsorized β values representing the proportion of methylation per CpG locus were subsequently used in statistical analyses.

### RNA-seq data collection

Whole blood samples were collected in PAXgene tubes (PreAnalytiX, Hombrechtikon, Switzerland). Isolation of total RNA was performed according to standard protocols (Asuragen, Inc., City, State) using the PAXgene Blood RNA System Kit as described previously.^16,17^ RNA-seq was performed in accordance with TOPMed protocols (University of Washington Northwest Genomics Center), and the mRNA-seq library was generated using the Illumina TruSeq (Illumina Inc., San Diego, CA). Stranded mRNA kit and sequencing was performed using the Illumina NovaSeq system (Illumina Inc., San Diego, CA). Gene expression was evaluated using RNA SeQC version 2.3.3^18^ and transcript expression was evaluated using RSEM version 1.3.1.^19^

### RNA-seq data pre-processing

Gene expression data were normalized using the trimmed mean of M-values (TMM) approach in the R *edgeR* package.^20^ A log2 transformation was applied to the TMM-normalized values after addition of 1 unit to avoid taking log of zero. This normalized gene expression value was residualized to account for batch effects, RNA concentration, and RNA integrity number.

### Statistical methods: eQTM analysis

We identified PCs from the residualized DNA methylation and gene expression data and performed an extensive search of the number of PCs to optimize the cross-validated replication of the CpG-transcript pairs. This study identified ten PCs for the DNA methylation data and five PCs for the gene expression data.

We then calculated the association between gene-level CpG sites and gene transcripts to identify significant eQTM CpG-transcript pairs. For each CpG-transcript pair, residualized gene expression was modeled as the outcome with residualized DNA methylation β values as the primary explanatory variable. These models were adjusted for age, sex, white blood cell count, blood cell fraction,^21^ platelet count, five gene expression PCs, and ten DNA methylation PCs. *Cis* gene-level CpG-transcript pairs were defined as those where the CpG site and the eGene (i.e., the gene transcript associated with the CpG) were within 1Mb kb of one another and were deemed statistically significant at a Bonferroni-corrected p-value of 1E-7.^22^ *Trans* pairs were defined as those where the CpG site and the eGene were more than 1Mb apart. While an estimated Bonferroni p-value cutoff of 1E-7/50,000 gene transcripts was calculated, we deemed *trans* pairs as statistically significant at a stricter p-value of 1E-14 to reduce noise and false positives based on an examination of the genomic positions of CpG sites and gene transcripts. This p-value threshold optimized the tradeoff between the replication rate and the number of significant pairs. Results were annotated to the Illumina CpG reference database^23^ and to GENCODE 30.

### Replication of eQTM CpG-transcript pairs

A recent eQTM analysis of nasal epithelial cells by Kim et al.^1^ identified 16,867 significant CpG-transcript pairs using the Illumina HumanMethylation450K methylation data and RNA-seq gene expression data. We mapped 8,481 CpG sites and 3,331 gene transcripts to those included in our eQTM analyses. Of these, we then identified CpG-transcript pairs that were also statistically significant in our eQTM results at p≤1E-7 for *cis* pairs and p≤1E-14 for *trans* pairs.

### Gene ontology

We evaluated gene ontology terms to identify specific biological and cellular pathways that were enriched among the top 1,000 unique *cis* and *trans* eGenes.^24,25^ Entrez gene IDs for the eGenes were included in these analyses, and we used the function “goana” in the R package *limma*^26^ to identify biological, cellular, and molecular pathways enriched in the eGenes. Statistically significant pathways were identified at a false discovery rate (FDR) ≤ 0.05.

### GWAS enrichment of clinical phenotypes associated with CpG-transcript pairs

We examined the overlap of the significant eQTM CpG-transcript pairs with gene-trait associations in the National Human Genome Research Institute (NHGRI)-European Bioinformatics Institute (EBI) GWAS catalog.^27^ The GWAS catalog included associations of 243,587 unique SNPs with 2,960 unique clinical traits at p ≤ 5E-8; SNPs were mapped to the gene(s) in which they were located. We compiled a list of unique gene transcripts that were part of statistically significant CpG-transcript pairs, as well as the associations of those genes with clinical phenotypes in the GWAS catalog. We subsequently used a one-sided Fisher’s exact test to evaluate enrichment for each gene with each of the 2,960 GWAS traits.

### Applications of eQTM resource: mediation and colocalization of cardiometabolic traits

To illustrate the application of the eQTM resource we generated, we provide examples from the associations of significant *cis* CpG-transcript pairs with three cardiovascular risk factor traits: serum triglycerides, fasting blood glucose, and body mass index (BMI). Specifically, we first evaluated the mediated effect of DNA methylation on the clinical phenotype through the expression of the linked eGene. Mediation analysis was performed using the function “mediate” in the R package *mediation* (version 4.5.0)^28^ to obtain total, direct, and mediated effects. This analysis provided the proportion of the total effect of the DNA methylation on the clinical phenotype mediated through gene expression. Results were divided by a factor of ten to obtain the effects associated with a 10% increase in DNA methylation of a CpG.

To identify candidate CpG-transcript pairs to include in mediation analysis, we identified 1) the overlap of significant *cis* CpG sites with published CpG-trait associations from the MRC Integrative Epidemiology Unit (IEU) EWAS catalog (http://www.ewascatalog.org/),^29^ and 2) significant results from *de novo* transcriptome-wide association studies (TWAS) of RNA-seq gene expression data and log-transformed serum triglycerides, fasting blood glucose, and BMI in the FHS. RNA-seq gene expression data for TWAS were residualized after adjustment for technical covariates and pedigree. All TWAS were then adjusted for age, sex, smoking status, alcohol use, white blood cell count, and predicted blood cell fraction.^21^ The TWAS of serum log triglycerides was additionally adjusted for BMI and use of lipid medications, and the TWAS of fasting glucose was additionally adjusted for BMI and diabetes status. Diabetes status was defined as a fasting glucose concentration ≥ 126 mg/dL or use of diabetes medications.

We conducted a Bayesian colocalization analysis to evaluate whether the *cis*-CpG-transcript eQTM pairs associated with serum log triglycerides, fasting blood glucose, and BMI were regulated by shared genetic variants. The MRC IEU EWAS catalog was used to identify CpG sites associated with each of the clinical phenotypes of interest, and three sources of data were used for colocalization analysis **(Figure 2)**. For the CpG-transcript pairs, we used FHS *cis* methylation quantitative loci (mQTL) and *cis* expression quantitative loci (eQTL) results to identify overlapping single nucleotide polymorphisms (SNPs) associated with the CpG and gene transcript, respectively. We then evaluated the overlap of *cis-*SNPs associated with CpG sites (mQTL) and *cis-*SNPs associated with gene transcripts (eQTL) with SNPs associated with serum log triglycerides,^30^ fasting blood glucose,^31^ and BMI^32^ in published genome-wide association studies (GWAS). We used the R package *coloc* (version 5.1.0)^33^ to quantify the probabilities (probability of colocalization H4) that a single genetic variant was associated with both DNA methylation and gene expression (i.e., colocalization of the mQTL and eQTL SNPs) and that a single genetic variant was associated with both DNA methylation and the clinical trait of interest (i.e., colocalization of the mQTL and GWAS SNPs).

**Figure 2.**
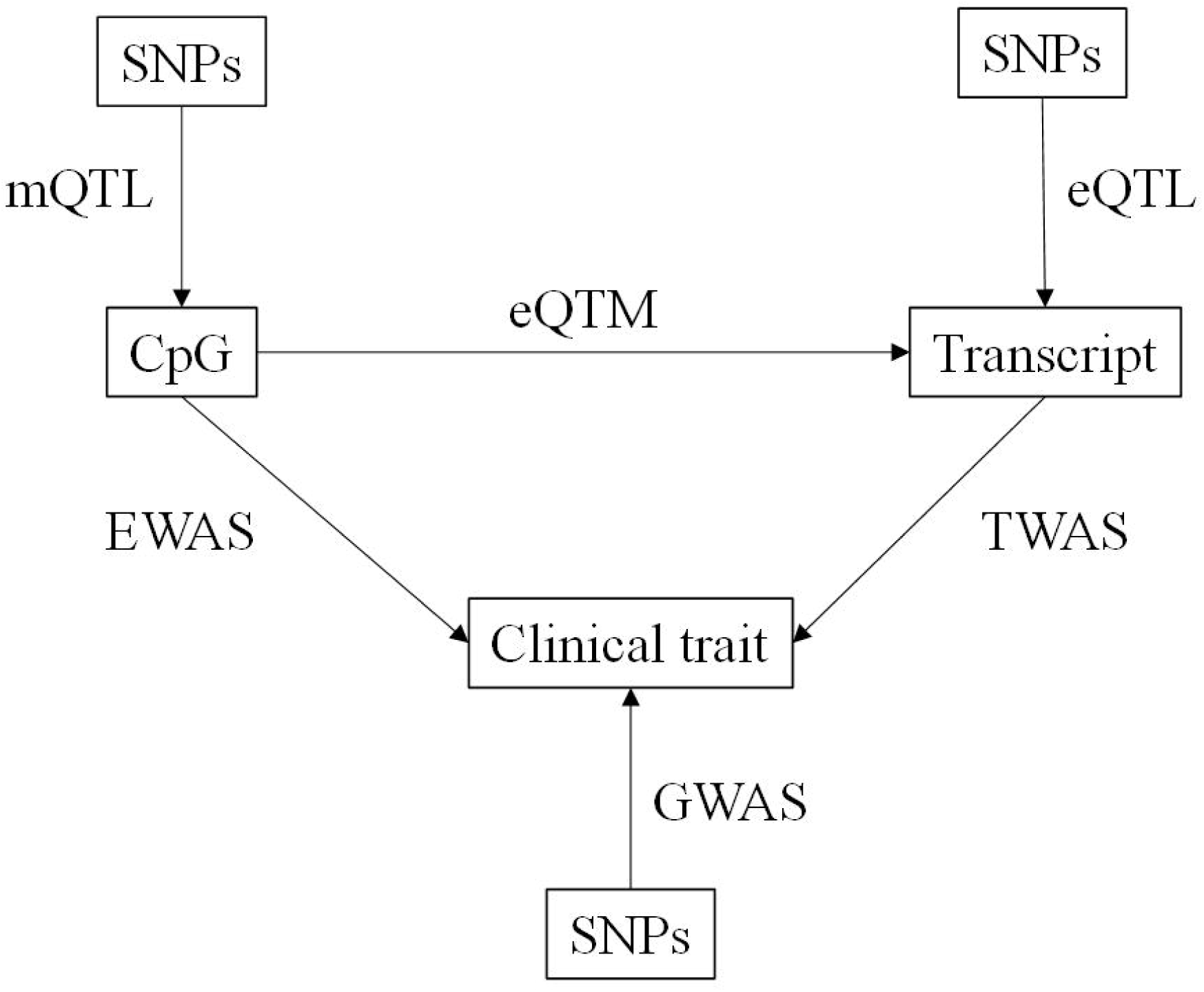
Overview of approach to combining eQTM results with other data sources for clinical and translational applications.

## Results

### Participant characteristics

The discovery sample included 2,115 participants (48% women) with an average age of 54 ± 15 years (**Table 1**). The average BMI was 28.0 ± 5.6 kg/m^2^, while the average serum triglycerides concentration was 114 ± 78 mg/dL. Average fasting blood glucose concentration was 100 ± 21 mg/dL.

**Table 1.**
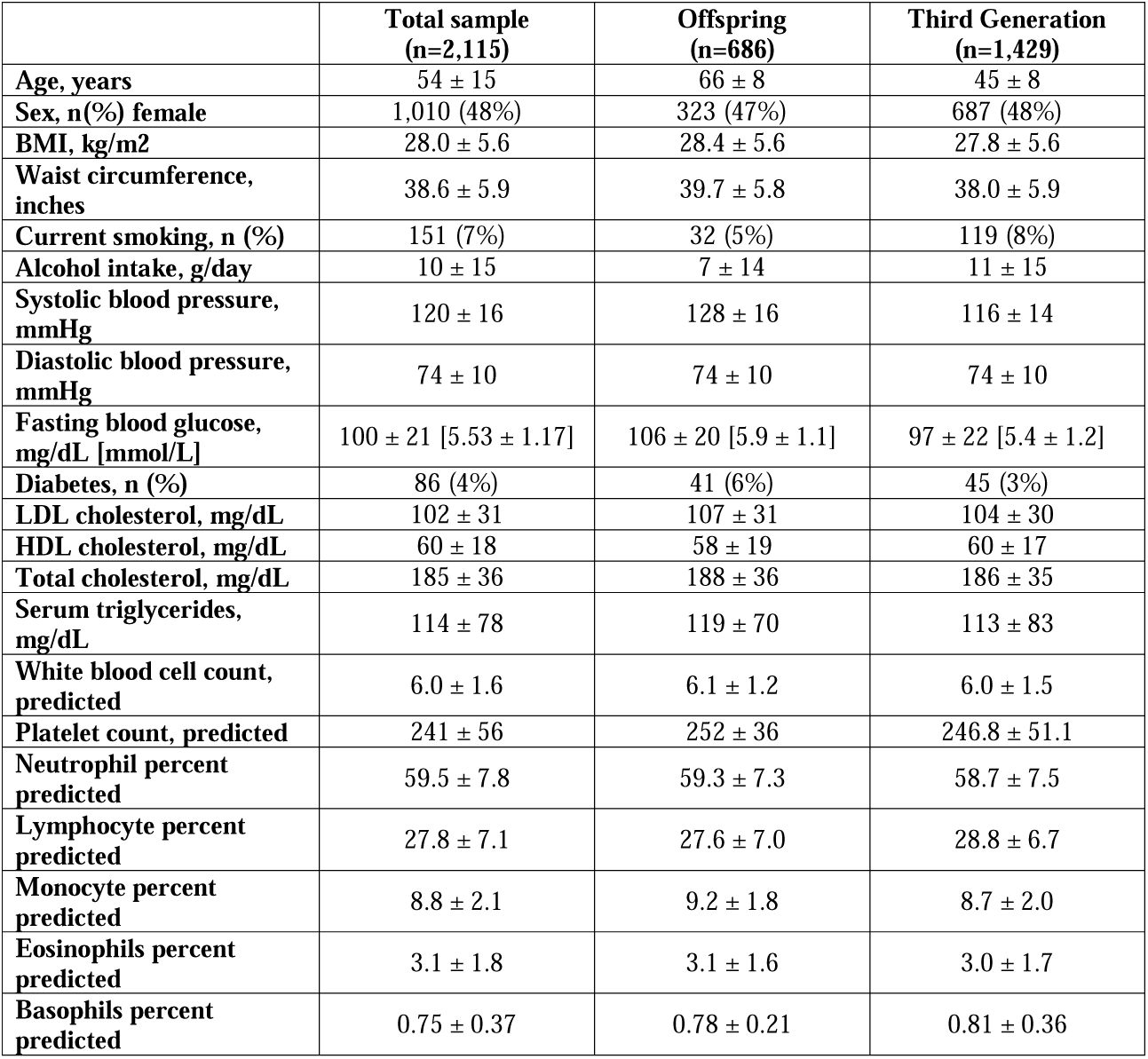
Characteristics of FHS participants.

|  | **Total sample**<br>**(n=2,115)** | **Offspring**<br>**(n=686)** | **Third Generation**<br>**(n=1,429)** |
| --- | --- | --- | --- |
| Age, years | 54 ± 15 | 66 ± 8 | 45 ± 8 |
| Sex, n(%) female | 1,010 (48%) | 323 (47%) | 687 (48%) |
| BMI, kg/m<sup>2</sup> | 28.0 ± 5.6 | 28.4 ± 5.6 | 27.8 ± 5.6 |
| Waist circumference, inches | 38.6 ± 5.9 | 39.7 ± 5.8 | 38.0 ± 5.9 |
| Current smoking, n (%) | 151 (7%) | 32 (5%) | 119 (8%) |
| Alcohol intake, g/day | 10 ± 15 | 7 ± 14 | 11 ± 15 |
| Systolic blood pressure, mmHg | 120 ± 16 | 128 ± 16 | 116 ± 14 |
| Diastolic blood pressure, mmHg | 74 ± 10 | 74 ± 10 | 74 ± 10 |
| Fasting blood glucose, mg/dL [mmol/L] | 100 ± 21 [5.53 ± 1.17] | 106 ± 20 [5.9 ± 1.1] | 97 ± 22 [5.4 ± 1.2] |
| Diabetes, n (%) | 86 (4%) | 41 (6%) | 45 (3%) |
| LDL cholesterol, mg/dL | 102 ± 31 | 107 ± 31 | 104 ± 30 |
| HDL cholesterol, mg/dL | 60 ± 18 | 58 ± 19 | 60 ± 17 |
| Total cholesterol, mg/dL | 185 ± 36 | 188 ± 36 | 186 ± 35 |
| Serum triglycerides, mg/dL | 114 ± 78 | 119 ± 70 | 113 ± 83 |
| White blood cell count, predicted | 6.0 ± 1.6 | 6.1 ± 1.2 | 6.0 ± 1.5 |
| Platelet count, predicted | 241 ± 56 | 252 ± 36 | 246.8 ± 51.1 |
| Neutrophil percent predicted | 59.5 ± 7.8 | 59.3 ± 7.3 | 58.7 ± 7.5 |
| Lymphocyte percent predicted | 27.8 ± 7.1 | 27.6 ± 7.0 | 28.8 ± 6.7 |
| Monocyte percent predicted | 8.8 ± 2.1 | 9.2 ± 1.8 | 8.7 ± 2.0 |
| Eosinophils percent predicted | 3.1 ± 1.8 | 3.1 ± 1.6 | 3.0 ± 1.7 |
| Basophils percent predicted | 0.75 ± 0.37 | 0.78 ± 0.21 | 0.81 ± 0.36 |

### Overview of eQTM results

We identified 70,047 significant *cis* CpG-transcript pairs (33,385 unique CpGs; 8,534 unique eGenes) at p≤1E-7. The majority (49,280; 70%) of the significant *cis* CpG-transcript pairs involved transcripts of protein-coding genes, while seven percent (4,971) were annotated to lncRNAs. Moreover, the 8,534 unique eGenes corresponded to 6,693 unique lead CpGs. DNA methylation of the lead CpG site was associated with increased *cis* expression of 2,909 (34%) eGenes, and with decreased *cis* expression of 5,625 (66%) eGenes.

We additionally identified 246,667 significant *trans* CpG-transcript pairs (12,637 unique CpGs, 4,300 unique eGenes) at p≤1E-14. The majority (210,944; 86%) of the significant *trans* CpG-transcript pairs involved transcripts of protein-coding genes, and 11,271 (5%) involved expression of lncRNAs. The 4,300 unique eGenes identified corresponded with 1,139 lead CpGs. DNA methylation of the lead CpG site was associated with increased *trans* expression of 1,566 (36%) eGenes, and with decreased *trans* expression of 2,734 (64%) eGenes.

To conduct internal validation, we divided our total sample into two equal-sized subsets: the first included all participants who had DNA methylation assayed by the 450K platform (discovery n=1,045), and the second included participants who had DNA methylation assayed by the EPIC platform (validation n=1,070). This internal validation analysis identified 40,148 significant *cis* CpG-transcript pairs in the discovery 450K samples at p≤1E-7. Of these, 79% (31,840 CpG-transcript pairs) were significant in the validation EPIC platform at a Bonferroni-corrected p-value≤0.05/40,148. We also identified 241,323 *trans* CpG-transcript pairs in the discovery sample at p≤1E-14, and of these, 31% (74,405 CpG-transcript pairs) were significant in the validation EPIC platform at a Bonferroni-corrected p-value≤0.05/241,323.

### External Replication

We identified 15,158 CpG-transcript pairs (8,481 CpGs and 3,331 transcripts) in the Kim et al.^1^ study, which we compared with our significant *cis* and *trans* eQTM results. Of these, 1,192 of 2,703 (44%) *cis* CpG-transcript pairs replicated in our study at p≤1E-7 with matching effect directionality. Additionally, 41 of 6,165 (0.67%) *trans* CpG-transcript pairs identified in Kim et al. replicated in our study at p≤1E-14 with matching effect directionality.

### Gene ontology

The top 1,000 unique *cis* eGenes were enriched in 33 pathways related to cell signaling and adhesion (**Supplemental Table 1**). The top 1,000 unique *trans* eGenes were enriched in 582 pathways related to the activation of the immune response **(Supplemental Table 2)**.

### GWAS enrichment

Using lead eQTL genetic variants for eQTM transcripts in enrichment analysis, the *cis* eGenes were enriched for genes associated with 1,208 traits in the GWAS catalog at an enrichment FDR ≤ 0.05 (**Supplemental Table 3**). The *trans* eGenes were enriched for genes associated with 1,191 traits in the GWAS catalog at an enrichment FDR ≤ 0.05 (**Supplemental Table 4**).

### eQTM CpG-transcript pairs in mediation analysis of cardiometabolic traits

The strategy of aligning trait-associated CpGs with corresponding trait-associated transcripts identified 14 CpG-transcript pairs associated with serum triglycerides, 36 associated with fasting glucose, and 153 associated with BMI. These pairs and their corresponding traits were then tested for the following mediation effects: 1) the total effect of DNA methylation on the cardiometabolic trait, 2) the direct effect of DNA methylation on the cardiometabolic trait independent of gene transcript expression, and 3) the mediated effect of DNA methylation on the cardiometabolic trait through gene transcript expression. Results are summarized in **Supplemental Table 5**. As an example, **Figure 3** illustrates significant mediation of the effects of DNA methylation at the CpG site cg11024682 on serum triglycerides, fasting blood glucose, and BMI via expression of the *SREBF1* gene. The proportion of the association between DNA methylation and the three cardiometabolic traits mediated by *SREBF1* expression ranged from 0.16 to 0.36.

**Figure 3.**
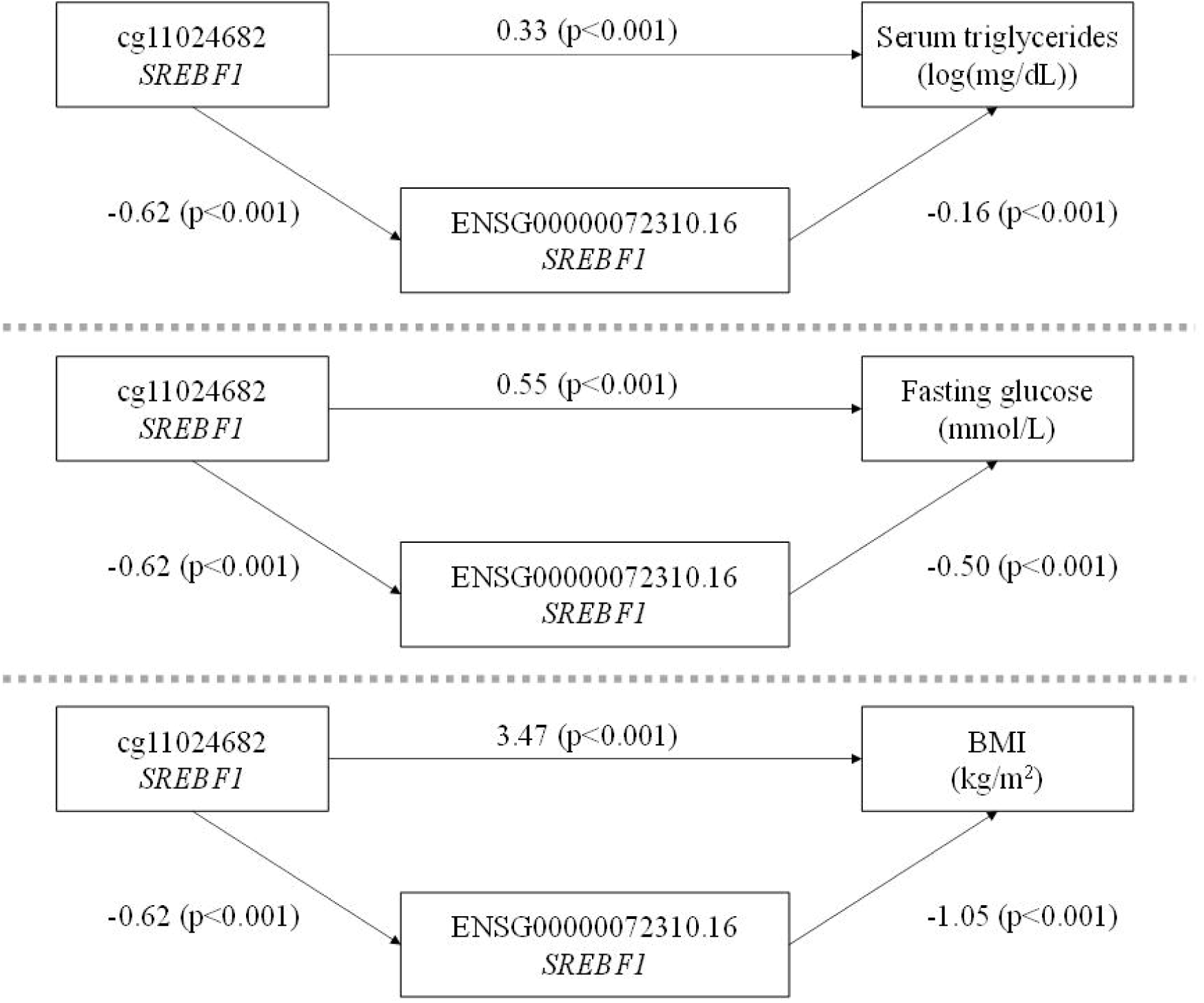
Pathways showing *cis* mediated effects of DNA methylation at cg11024682 on serum triglycerides, fasting blood glucose, and BMI through the expression of *SREBF1*. The effects of DNA methylation at the CpG site cg11024682 on all three cardiometabolic traits of interest are mediated through *cis* effects on *SREBF1* expression. Coefficients presented in the figure represent the change in gene expression associated with a 10% increase in DNA methylation; the change in serum triglycerides (log(mg/dL)), fasting blood glucose (mmol/L), or BMI (kg/m^2^) associated with a 10% increase in DNA methylation; or the change in serum triglycerides (log(mg/dL)), fasting blood glucose (mmol/L), or BMI (kg/m^2^) associated with a 1-unit increase in gene expression.

Gene expression mediated the effect of DNA methylation on serum triglycerides (log-transformed) for 10 of the 14 CpG-transcript pairs. Significant mediation of DNA methylation on gene expression was observed for the expression of *RNASET2, SREBF1, ABCG1*, and *TOM1L2* at p≤0.05.

Gene transcript expression significantly mediated the effect of DNA methylation on fasting blood glucose for 24 of the 36 CpG-transcript pairs, of which six had direct and mediated effects in opposite directions (e.g., a negative direct effect and a positive mediated effect). The greatest proportions of the CpG-fasting glucose associations mediated by gene expression (p<2E-16 for all) were observed for the expression of *KLRF1* (proportion mediated =0.87), *TRAPPC2B* (proportion mediated=0.82), *NKIRAS2* (proportion mediated=0.74), and *RRP12* (proportion mediated=0.52).

Finally, gene transcript expression significantly mediated the effect of DNA methylation on BMI for 117 of the 153 CpG-transcript pairs, of which 55 had direct and mediated effects in opposite directions. For the CpG-transcript pairs where the direct and mediated effects were in the same direction, the greatest proportions of the CpG-BMI associations mediated by gene expression (p<2E-16 for all) were observed for the expression of *TSPYL1* (proportion mediated=0.98), *PLD3* (proportion mediated=0.94), and *FAS* (proportion mediated=0.80). Additionally, two CpG-transcript pairs showed significant mediation of the CpG-BMI association through expression of lncRNA genes *BX284668*.*5* and *LINC00996*.

### eQTM CpG-transcript pairs in colocalization analysis of cardiometabolic traits

Among significant serum triglycerides-associated CpG sites identified in the MRC IEU EWAS catalog, 15 overlapped with CpGs from significant *cis* CpG-transcript pairs where the CpG and gene transcript also shared at least one SNP (i.e., the eQTL variant for the transcript matched the mQTL variant for its paired CpG). Colocalization results for the 15 *cis* eQTM CpG-transcript pairs, which included 12 unique CpG sites associated with 4,876 significant *cis-*mQTL variants and 11 unique gene transcripts associated with 6,327 significant *cis*-eQTL variants, are presented in **Supplemental Table 6**. The probability of colocalization H4 ≥ 80% was observed between the CpG and transcript SNPs for 10 eQTM CpG-transcript pairs. The SNP rs7215055 was associated with serum triglycerides^30^ and was also associated with methylation of CpG cg11024682 and expression of gene *ATPAF2* (probability of colocalization H4=100%).

Among significant fasting glucose-associated CpG sites identified in the MRC IEU EWAS catalog, 138 overlapped with CpGs from significant *cis-*CpG-transcript pairs where the CpG and gene transcript also shared at least one SNP. Colocalization results for the 138 *cis-*eQTM CpG-transcript pairs, which included 109 unique CpG sites associated with 38,758 significant *cis-*mQTL variants and 172 unique gene transcripts associated with 100,546 significant *cis-*eQTL variants, are presented in **Supplemental Table 7**. Probability of colocalization ≥ 80% was observed between the CpG and transcript SNPs for 51 eQTM CpG-transcript pairs. There was no overlap of SNPs identified in a GWAS of fasting glucose^31^ with both mQTL and eQTL variants for CpG-transcript pairs, thus further colocalization analyses were not carried out for this trait.

Finally, among significant BMI-associated CpG sites identified in the MRC IEU EWAS catalog, 207 overlapped with CpGs from significant *cis* CpG-transcript pairs where the CpG and gene transcript also shared at least one SNP. Colocalization results for the 207 *cis-*eQTM CpG-transcript pairs, which included 134 unique CpG sites associated with 82,823 significant *cis-*mQTL variants and 182 unique gene transcripts associated with 232,511 significant *cis-*eQTL variants, are presented in **Supplemental Table 8**. Probability of colocalization ≥ 80% was observed between the CpG and transcript for 67 eQTM CpG-transcript pairs. The SNPs rs3817334, rs10838738, and rs1064608, which were associated with BMI,^32^ were also mQTL genetic variants associated with the CpG cg17580616 and eQTL genetic variants associated with the gene *ACP2* (probability of colocalization H4=100%).

## Discussion

Our study of DNA methylation in relation to gene expression in 2,115 FHS participants identified over 70,000 significant *cis* CpG-transcript pairs and over 246,000 significant *trans* CpG-transcript pairs. We generated a comprehensive database of *cis* and *trans* eQTM CpG-transcript pairs and will make this resource available publicly to investigate disease mechanisms, pathways, and processes. We previously evaluated eQTM CpG-transcript pairs in the FHS using the Illumina 450K DNA methylation array in conjunction with array-based gene expression data.^7^ The present study expands on this prior work by incorporating more extensive DNA methylation profiling (Illumina EPIC DNA methylation array) and RNA-seq expression, resulting in a powerful resource for capturing associations between DNA methylation and gene expression. Not only did we identify four times as many *cis*-eQTM CpG-transcript pairs (70,047 vs. 16,416) and 24% more *trans*-pairs (246,667 vs. 198,960) than we previously identified using array-based gene expression data,^7^ but we also provide eQTM CpG-transcript pairs for the expression of lncRNAs.

DNA methylation can alter the regulation of gene transcription in multiple ways, such as aiding in the recruitment of proteins involved in increasing gene expression or inhibiting transcription factor binding to a specific DNA sequence. At promoter sites, DNA methylation generally precludes transcription directly by blocking the binding of transcriptional activators or indirectly by recruiting methyl-binding proteins and co-repressor complexes. In this study, we used eQTM analysis to identify numerous CpG sites that, when methylated, are associated with *cis* and *trans* eGene expression. To showcase the potential clinical utility of our findings, we evaluated the role of our eQTM resource when applied to the analysis of three cardiometabolic risk factors – serum triglycerides, fasting blood glucose, and BMI – in mediation and colocalization analyses. We illustrate specific examples of how the eQTM resource can be used to suggest molecular mechanisms linking DNA methylation to clinical phenotypes: mediation analyses identified putative pathways by which DNA methylation is associated with clinical traits in EWAS, and colocalization analysis suggested shared genetic regulation of CpGs, transcripts, and clinical traits. For example, the results of our mediation analysis showed that expression of the genes *ABCG1* and *SREBF1* mediates the association between DNA methylation and all three clinical traits.

While the eQTM resource we created is robust, several limitations must be noted. First, FHS is an observational study consisting of participants of predominantly European ancestry. Results may not be generalizable to different races or ethnicities, and these results have not been validated in independent external cohorts. Second, while the use of DNA methylation and gene expression data derived from whole blood samples makes our methods more accessible and more easily replicated in other study cohorts, evaluation of CpG-transcript pairs in other tissues may be more relevant to specific disease processes. Finally, this study does not examine the effect of environmental factors on these associations; lifestyle factors such as cigarette smoking are known to affect DNA methylation and gene expression.^7,34–36^ Future eQTM research may incorporate environmental data to better understand the epigenetic effects of environmental exposures.

## Conclusions

We created a powerful eQTM resource that leverage DNA methylation and RNA-seq data to characterize associations of DNA methylation with gene expression. A comprehensive summary data set will be posted to the National Heart, Lung, and Blood Institute’s BioData Catalyst site. We provide proof of concept that eQTM resources can be leveraged to explore molecular mechanisms of disease.

## Data Availability

All data produced in the present study are available upon reasonable request to the authors.

## Acknowledgements

This work utilized the computational resources of the NIH HPC Biowulf cluster. (http://hpc.nih.gov). Data on glycemic traits have been contributed by MAGIC investigators and have been downloaded from www.magicinvestigators.org.

## Disclaimer

The views and opinions expressed in this manuscript are those of the authors and do not necessarily represent the views of the National Heart, Lung, and Blood Institute, the National Institutes of Health, or the U.S. Department of Health and Human Services.

## References

1. Kim S, Forno E, Zhang R, et al. Expression Quantitative Trait Methylation Analysis Reveals Methylomic Associations With Gene Expression in Childhood Asthma. Chest. 2020;158(5):1841–1856. doi:https://doi.org/10.1016/j.chest.2020.05.601

2. Samblas M, Milagro FI, Martínez A. DNA methylation markers in obesity, metabolic syndrome, and weight loss. Epigenetics. 2019;14(5):421–444. doi:10.1080/15592294.2019.1595297

3. Płatek T, Polus A, Góralska J, et al. DNA methylation microarrays identify epigenetically regulated lipid related genes in obese patients with hypercholesterolemia. Mol Med. 2020;26(1):93. doi:10.1186/s10020-020-00220-z

4. Liu J, Carnero-Montoro E, van Dongen J, et al. An integrative cross-omics analysis of DNA methylation sites of glucose and insulin homeostasis. Nat Commun. 2019;10(1):2581. doi:10.1038/s41467-019-10487-4

5. Richard MA, Huan T, Ligthart S, et al. DNA Methylation Analysis Identifies Loci for Blood Pressure Regulation. Am J Hum Genet. 2017;101(6):888–902. doi:10.1016/j.ajhg.2017.09.028

6. Myte R, Sundkvist A, Van Guelpen B, Harlid S. Circulating levels of inflammatory markers and DNA methylation, an analysis of repeated samples from a population based cohort. Epigenetics. 2019;14(7):649–659. doi:10.1080/15592294.2019.1603962

7. Yao C, Joehanes R, Wilson R, et al. Epigenome-wide association study of whole blood gene expression in Framingham Heart Study participants provides molecular insight into the potential role of CHRNA5 in cigarette smoking-related lung diseases. Clin Epigenetics. 2021;13(1):60. doi:10.1186/s13148-021-01041-5

8. Sharma NK, Comeau ME, Montoya D, et al. Integrative Analysis of Glucometabolic Traits, Adipose Tissue DNA Methylation, and Gene Expression Identifies Epigenetic Regulatory Mechanisms of Insulin Resistance and Obesity in African Americans. Diabetes. 2020;69(12):2779–2793. doi:10.2337/db20-0117

9. Maas SCE, Mens MMJ, Kühnel B, et al. Smoking-related changes in DNA methylation and gene expression are associated with cardio-metabolic traits. Clin Epigenetics. 2020;12(1):157. doi:10.1186/s13148-020-00951-0

10. Rao MS, Van Vleet TR, Ciurlionis R, et al. Comparison of RNA-Seq and Microarray Gene Expression Platforms for the Toxicogenomic Evaluation of Liver From Short-Term Rat Toxicity Studies. Front Genet. 2019;9. https://www.frontiersin.org/article/10.3389/fgene.2018.00636

11. Ismail N, Abdullah N, Abdul Murad NA, Jamal R, Sulaiman SA. Long Non-Coding RNAs (lncRNAs) in Cardiovascular Disease Complication of Type 2 Diabetes. Diagnostics (Basel, Switzerland). 2021;11(1):145. doi:10.3390/diagnostics11010145

12. Maass PG, Luft FC, Bähring S. Long non-coding RNA in health and disease. J Mol Med (Berl). 2014;92(4):337–346. doi:10.1007/s00109-014-1131-8

13. Kannel WB, Feinleib M, McNamara PM, Garrison RJ, Castelli WP. An investigation of coronary heart disease in families: the Framinham Offspring Study. Am J Epidemiol. 1979;110(3):281–290. doi:10.1093/oxfordjournals.aje.a112813

14. Tsao CW, Vasan RS. Cohort Profile: The Framingham Heart Study (FHS): overview of milestones in cardiovascular epidemiology. Int J Epidemiol. 2015;44(6):1800–1813. doi:10.1093/ije/dyv337

15. Pidsley R, Y Wong CC, Volta M, Lunnon K, Mill J, Schalkwyk LC. A data-driven approach to preprocessing Illumina 450K methylation array data. BMC Genomics. 2013;14(1):293. doi:10.1186/1471-2164-14-293

16. Joehanes R, Johnson AD, Barb JJ, et al. Gene expression analysis of whole blood, peripheral blood mononuclear cells, and lymphoblastoid cell lines from the Framingham Heart Study. Physiol Genomics. 2011;44(1):59–75. doi:10.1152/physiolgenomics.00130.2011

17. Joehanes R, Ying S, Huan T, et al. Gene expression signatures of coronary heart disease. Arterioscler Thromb Vasc Biol. 2013;33(6):1418–1426. doi:10.1161/ATVBAHA.112.301169

18. DeLuca DS, Levin JZ, Sivachenko A, et al. RNA-SeQC: RNA-seq metrics for quality control and process optimization. Bioinformatics. 2012;28(11):1530–1532. doi:10.1093/bioinformatics/bts196

19. Li B, Dewey CN. RSEM: accurate transcript quantification from RNA-Seq data with or without a reference genome. BMC Bioinformatics. 2011;12:323. doi:10.1186/1471-2105-12-323

20. Robinson MD, McCarthy DJ, Smyth GK. edgeR: a Bioconductor package for differential expression analysis of digital gene expression data. Bioinformatics. 2010;26(1):139–140. doi:10.1093/bioinformatics/btp616

21. Houseman EA, Accomando WP, Koestler DC, et al. DNA methylation arrays as surrogate measures of cell mixture distribution. BMC Bioinformatics. 2012;13:86. doi:10.1186/1471-2105-13-86

22. Saffari A, Silver MJ, Zavattari P, et al. Estimation of a significance threshold for epigenome-wide association studies. Genet Epidemiol. 2018;42(1):20–33. doi:10.1002/gepi.22086

23. Illumina I. Technical Note: Epigenetics - CpG Loci Identification.; 2010. https://www.illumina.com/content/dam/illumina-marketing/documents/products/technotes/technote_cpg_loci_identification.pdf

24. Ashburner M, Ball CA, Blake JA, et al. Gene ontology: tool for the unification of biology. The Gene Ontology Consortium. Nat Genet. 2000;25(1):25–29. doi:10.1038/75556

25. The Gene Ontology resource: enriching a GOld mine. Nucleic Acids Res. 2021;49(D1):D325–D334. doi:10.1093/nar/gkaa1113

26. Ritchie ME, Phipson B, Wu D, et al. limma powers differential expression analyses for RNA-sequencing and microarray studies. Nucleic Acids Res. 2015;43(7):e47–e47. doi:10.1093/nar/gkv007

27. Buniello A, MacArthur JAL, Cerezo M, et al. The NHGRI-EBI GWAS Catalog of published genome-wide association studies, targeted arrays and summary statistics 2019. Nucleic Acids Res. 2019;47(D1):D1005–D1012. doi:10.1093/nar/gky1120

28. Tingley D, Teppei H, Mit Y, Keele L, State P, Imai K. Mediation: R Package for Causal Mediation Analysis. J Stat Softw. 2014;59. doi:10.18637/jss.v059.i05

29. Battram T, Yousefi P, Crawford G, et al. The EWAS Catalog: a database of epigenome-wide association studies. OSF Prepr. Published online 2021:4. doi:10.31219/OSF.IO/837WN

30. Richardson TG, Sanderson E, Palmer TM, et al. Evaluating the relationship between circulating lipoprotein lipids and apolipoproteins with risk of coronary heart disease: A multivariable Mendelian randomisation analysis. PLoS Med. 2020;17(3):e1003062–e1003062. doi:10.1371/journal.pmed.1003062

31. Chen J, Spracklen CN, Marenne G, et al. The trans-ancestral genomic architecture of glycemic traits. Nat Genet. 2021;53(6):840–860. doi:10.1038/s41588-021-00852-9

32. Turcot V, Lu Y, Highland HM, et al. Protein-altering variants associated with body mass index implicate pathways that control energy intake and expenditure in obesity. Nat Genet. 2018;50(1):26–41. doi:10.1038/s41588-017-0011-x

33. Giambartolomei C, Vukcevic D, Schadt EE, et al. Bayesian test for colocalisation between pairs of genetic association studies using summary statistics. PLoS Genet. 2014;10(5):e1004383–e1004383. doi:10.1371/journal.pgen.1004383

34. Joehanes R, Just AC, Marioni RE, et al. Epigenetic Signatures of Cigarette Smoking. Circ Cardiovasc Genet. 2016;9(5):436–447. doi:10.1161/CIRCGENETICS.116.001506

35. Keshawarz A, Joehanes R, Guan W, et al. Longitudinal change in blood DNA epigenetic signature after smoking cessation. Epigenetics. Published online October 2021:1–12. doi:10.1080/15592294.2021.1985301

36. Huan T, Joehanes R, Schurmann C, et al. A whole-blood transcriptome meta-analysis identifies gene expression signatures of cigarette smoking. Hum Mol Genet. 2016;25(21):4611–4623. doi:10.1093/hmg/ddw288

